# SARS-CoV-2 infections in airway epithelial cells from smokers versus non-smokers

**DOI:** 10.1101/2021.04.28.21255760

**Authors:** Manel Essaidi-Laziosi, Giulia Torriani, Catia Alvarez, Laurent Kaiser, Isabella Eckerle

## Abstract

Whether smoking exacerbates Coronavirus disease 2019 is still debated. *Ex-vivo* Infection of reconstituted epithelial tissues from smoker versus non-smoker donors suggested comparable susceptibility to SARS-CoV-2 in epithelia from both groups.

---

Controversial conclusions have been drawn from epidemiological studies reporting both an increased and decreased risk of both Severe Acute Respiratory Syndrome Coronavirus 2 (SARS-CoV-2) infection and Coronavirus disease 2019 (COVID-19) exacerbation for smokers [1]. Although a synopsis of available evidence suggests that current smoking is associated with an increased disease severity and death rate in hospitalized COVID-19 patients [2], further investigations are still needed. Also, cumulative smoking exposure was recently reported as an independent risk factor for hospitalization and death due to COVID19 [3]. Pathophysiological mechanisms involved in a higher susceptibility to infection or disease could be multifactorial, as it is known that smoking leads to altered expression of inflammatory cytokines and impaired innate immunity in the mucosa, increased permeability of epithelial cells and impaired mucociliary clearance function, as well as epigenetic modifications within the epithelium of the respiratory tract. In particular, overexpression of Angiotensin Converting Enzyme II (ACE-2) in the respiratory tract, leading to more efficient entry of SARS-CoV-2 into cells, was hypothesized to increase risk of SARS-CoV-2 infection and COVID-19 [4-6].

*In vitro* reconstituted human primary airway epithelial cells constitute an appropriate surrogate model for studying the first steps of respiratory viral infections, closely mimicking the *in vivo* situation [7]. Such primary cells are available from healthy donors, but also from patients with pre-existing respiratory disorders, including cystic fibrosis, chronic obstructive pulmonary disease, asthma and also from individuals with a history of smoking.

In our study, we have investigated SARS-CoV-2 replication in reconstituted upper respiratory tract (URT) airway epithelial cells (AECs) derived from the bronchial epithelium from individuals with (n=3, 25 to 80 pack years), or without (n=3, no other pathologies) a history of smoking (**Table 1**). All epithelial tissues originating from smokers and non-smokers donors have been purchased from Epthelix SARL(https://www.epithelix.com/). Their production was conducted according to the Declaration of Helsinki on biomedical research (Hong Kong amendment, 1989), and the research protocol was approved by the local ethics committee.

To better recapitulate the *in vivo* situation, infections were performed in air-liquid interface (ALI) system, as previously described [8], and a SARS-CoV-2 stock was obtained from a clinical isolate collected during the first pandemic wave in March 2020 on AECs, to avoid any adaptation to cell culture during the isolation process (initial sequence registered at GISAID hCoV-19/Switzerland/GE-SNRCI-29943121/2020|EPI_ISL_414019|2020-02-27). Sequencing the initial patient specimen and the isolate did not reveal any mutations. Infection assays were performed at 37°C and a multiplicity of infection (MOI) of 0.01. Viral replication was assessed by quantitative real time PCR (RT-qPCR) from RNA extracted from apical tissue washes collected at days 2 and 4 post-infection (dpi). Compared to 3 hours post infection, SARS-CoV-2 replication was equally efficient at 2 (2.07E10, SD± 3.9E10 SARS-CoV-2 RNA copies/mL) and 4 dpi (1.26E11 SD ±1.82E+11 SARS-CoV-2 RNA copies/mL) in tissues from both smokers and non-smokers (**Figure 1A**). At day 4 post infection, interferon and ACE-2 inductions were determined by RT-qPCR from total RNA extracted from the cell lysates and represented as the fold change relative to non-infected tissues (figure 2). As expected [8], type III IFN induction in infected tissues (10E4 fold increase) is higher than type I (**Figure 1B**). Comparing smokers and non-smokers, similar expression levels of IFNs α, β and λ and ACE-2 were also observed in mock- (data not shown) and SARS-CoV-2-infected tissues at 4 dpi (Figure 1B). Altogether, our data showed similar *ex vivo* viral replication and host-response in SARS-CoV-2-infected tissues of the upper respiratory tract from smokers versus non-smoker donors, indicating comparable susceptibility to SARS-CoV-2 in bronchial epithelia from both groups.

**Figure 1.**
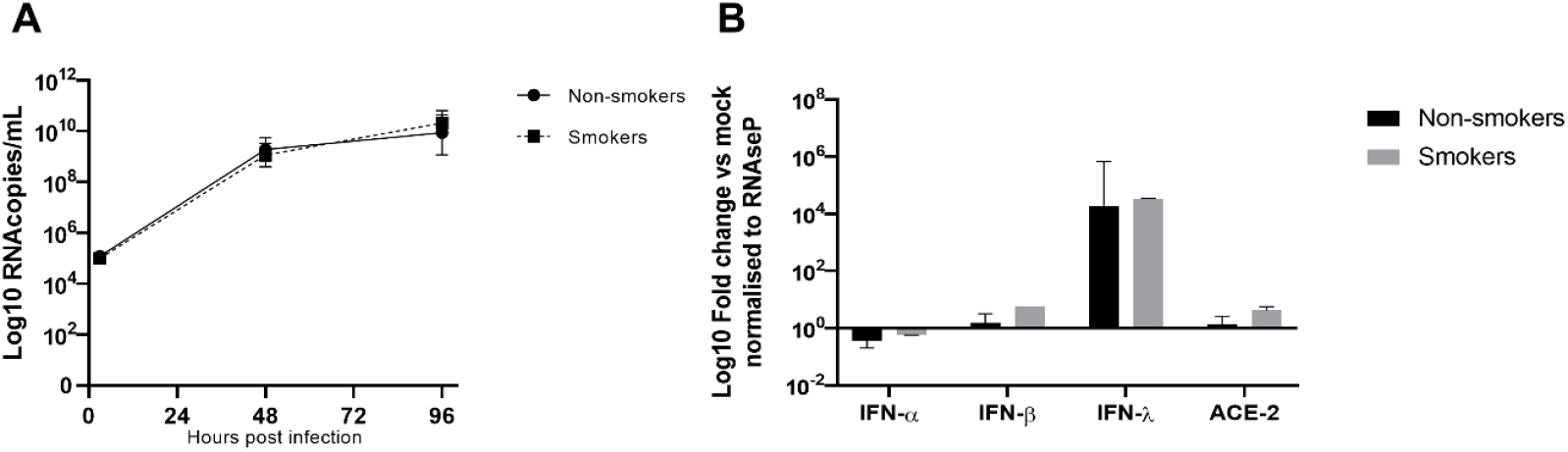
SARS-CoV2 infections in 3D tissues from non-smoker versus smoker donors. A. Virus replication. Virus production at the apical side of air-liquid interface culture (37°C, MOI=0.01) of reconstituted human airway epithelia[8] derived from non-smoker (N=3) versus smoker (N=3) patients were compared at days 2 and 4 pi by quantitative real time PCR targeting the E gene. Four tissues have been tested for each donor. B. Host response induction in SARS-CoV2 infected tissues. The expression of IFNs α, β and λ and ACE-2 receptor at day 4pi was semi-quantified by RT-qPCR on total RNA extracted from tissue lysates, normalized to housekeeping gene (RNAseP). Fold change of IFNs α, β and λ and ACE-2 inductions in infections relative to non-infected tissues from non-smoker (N=3) versus smoker (N=3) patients were compared. Four tissues were tested for each donor. Statistical significance calculated using the two-way ANOVA showed non significant differences in viral replication and host response in tissues from the two groups.

Although several studies have found an influence of smoking on increased ACE-2 expression mainly in the lower respiratory tract (LRT), with potentially higher susceptibility to, and increased replication of, SARS-CoV-2; no such effect was observed in our model on AECs from the URT, the primary entry site of SARS-CoV-2 [9].

In our highly relevant *ex vivo* model derived from the airway epithelium, similar susceptibility to viral infection and replication, as well as comparable host profiles, in both groups were observed. Thus ACE-2 expression, higher tissue-permissiveness to SARS-CoV-2, or differences in interferon response at the site of entry, are most likely not responsible for pronounced disease. Other factors like involvement of activated immune cells and adaptive immunity (lacking in our model), higher infection susceptibility of the LRT and the role of additional comorbidities associated with smoking could constitute risk factors for pronounced COVID-19. Further investigations of these mechanisms are still needed.

The possibility of reversible smoking-induced alterations being restored in our reconstituted tissues presents the main limitation of our model. Using ALI cultures from primary human non-smoker airway basal stem cells, Purkayastha A. *et al* recently showed that direct short-term exposure to cigarette smoke led to more severe SARS-CoV-2 disease due to the reduction of the immune response [10].

In conclusion, although no evidence of increased COVID-19 disease vulnerability in smokers was found, this work shows that, if such susceptibility exists, it would not involve early infection steps and is unlikely to be dependent on the response of the upper respiratory tract epithelium *per se*.

## Data Availability

Data available on request from the authors

## Abbreviations

SARS-CoV-2: Severe Acute Respiratory Syndrome Coronavirus 2
COVID119: coronavirus disease 2019
URT/LRT: upper/lower respiratory tract
AECs: airway epithelial cells
ALI: air-liquid interface
MOI: multiplicity of infection
ACE-2: Angiotensin Converting Enzyme II
IFN: Interferon

## Acknowledgments

We thank Pascale Sattonnet-Roche for excellent technical support and Erik Boehm for English proof reading (Geneva Centre for Emerging Viral Diseases, Geneva University Hospitals, Geneva, Switzerland).

